# The need of health policy perspective to protect Healthcare Workers during COVID-19 pandemic. A GRADE rapid review on the N95 respirators effectiveness

**DOI:** 10.1101/2020.04.06.20054841

**Authors:** Primiano Iannone, Greta Castellini, Daniela Coclite, Antonello Napoletano, Alice Fauci, Laura Iacorossi, Daniela D’Angelo, Cristina Renzi, Giuseppe La Torre, Claudio M. Mastroianni, Silvia Gianola

**Affiliations:** Centro Eccellenza Clinica, Qualità e Sicurezza delle Cure, Istituto Superiore di Sanità, Rome, Italy; IRCCS Istituto Ortopedico Galeazzi, Unit of Clinical Epidemiology, Milan, Italy; University College London-UCL, Institute of Epidemiology & Health Care, London, UK; Department of Public Health and Infectious Diseases, Sapienza University of Rome, Rome, Italy

**Keywords:** Coronavirus, COVID–19, SARS-CoV-2, surgical mask, N95 respirator, respiratory protective devices, respiratory tract infections, healthcare workers, prevention

## Abstract

Protecting Health Care Workers (HCWs) during routine care of suspected or confirmed COVID-19 patients is of paramount importance to halt the SARS-CoV-2 (Severe Acute Respiratory Syndrome-Coronavirus-2) pandemic. The WHO, ECDC and CDC have issued conflicting guidelines on the use of respiratory filters (N95) by HCWs. We searched PubMed, Embase and The Cochrane Library from the inception to March 21, 2020 to identify randomized controlled trials (RCTs) comparing N95 respirators versus surgical masks for prevention of COVID-19 or any other respiratory infection among HCWs. The grading of recommendations, assessment, development, and evaluation (GRADE) was used to evaluate the quality of evidence. Four RCTs involving 8736 HCWs were included. We did not find any trial specifically on prevention of COVID-19. However, wearing N95 respirators can prevent 73 more (95% CI 46-91) clinical respiratory infections per 1000 HCWs compared to surgical masks (2 RCTs; 2594 patients; low quality of evidence). A protective effect of N95 respirators in laboratory-confirmed bacterial colonization (RR= 0.41; 95%CI 0.28-0.61) was also found. A trend in favour of N95 respirators was observed in preventing laboratory-confirmed respiratory viral infections, laboratory-confirmed respiratory infection, and influenza like illness. We found no direct high quality evidence on whether N95 respirators are better than surgical masks for HCWs protection from SARS-CoV-2. However, low quality evidence suggests that N95 respirators protect HCWs from clinical respiratory infections. This finding should be contemplated to decide the best strategy to support the resilience of healthcare systems facing the potentially catastrophic SARS-CoV-2 pandemic.

## Introduction

The Severe Acute Respiratory Syndrome-Coronavirus-2 (SARS-CoV-2) outbreak emerged in China in December 2019 and it was recognised as a pandemic by the World Health Organization (WHO) on 11 March[1]. As of 30 March 2020, a total of 693,224 cases and 33,106 deaths have been reported worldwide[2]. Nosocomial spread and infection of healthcare workers (HCWs) are a major concern. In Italy HCWs are paying a heavy price in addition to their professional and humanitarian efforts, with 8956 cases (more than 9% of total Italian cases[3]) and 63 deaths[4] among physicians. Protecting HCWs from SARS-CoV-2 is therefore of great importance for individual HCW and for their role in fighting this devastating pandemic effectively. Claims of insufficient protection of HCWs by personal protective equipment, in particular with regards to the use of surgical masks, have fuelled the scientific and social media debate in Italy. In fact, except for aerosol generating procedures requiring higher level of respiratory protection with filtering respirators, WHO considers surgical masks adequate for the routine care of coronavirus disease 2019 (COVID-19) patients [5]. Instead, the Centers for Disease Control (CDC) and the European Center for Disease Control guidelines (ECDC) have a more cautious approach, acknowledging that the exact role of airborne (aerosol) route in the transmission of SARS-CoV-2 is still largely unknown[6, 7]. The direct evidence supporting the WHO guidelines is based on very few case reports on the absence of SARS-CoV-2 in air samples taken in highly protected environments where a rapid dilution of aerosols occurs, the absence of infection of HCWs exposed for a limited time or limited viral loads, or on modelling of epidemiologic patterns of transmission[8-11]. In contrast, the airborne (aerosol) opportunistic route of transmission has been documented for SARS and MERS caused by closely related coronaviruses responsible of severe nosocomial infections among HCWs. Aerorsol filtering respirators were consequently recommended for SARS during 2002–03 outbreak [12]. It is worth remembering that Canadian Health authorities modified their earlier recommendations in favour of a more strict respiratory protection after the deaths of several HCWs [13]. The presence of SARS-CoV-2 in aerosols has been documented in experimental [14] and real life conditions in crowded, poorly ventilated hospital areas unrelated to aerosol generating procedures [15]. Also, spontaneous cough generates aerosols, not only droplets [16, 17] and COVID-19 patients may infect HCWs in this way, especially if they are unable to wear facemasks due to hypoxia and need of oxygen therapy. Moreover, none of the above mentioned guidelines adopted the suggested Grading of Recommendations Assessment, Development and Evaluation (GRADE) approach for making public health policy guidelines and they did not explicitly consider the potentially catastrophic consequences of deferring the recommendation of N95 for HCWs while awaiting more robust evidence.

There are already some systematic reviews addressing the role of N95 respirators in protecting HCWs, offering however biased estimates of the effect [18-20]. We therefore undertook a systematic review with a different perspective and methodology, given the exceptional disease burden expected from this pandemic [21], the central role of protecting HCWs and the need of a careful definition of the outcomes, which are critical for unbiased public health policy decisions [22]. Indeed strengthening the preparedness and resiliency of health care systems to this pandemic crisis occurs not only avoiding SARS-CoV-2 infection but also preventing any HCW respiratory infection causing absenteeism from work. We therefore conducted a systematic review aimed at assessing the efficacy of N95 respirators versus surgical masks for the prevention of respiratory tract infections transmission among HCWs. The evidence from the review can then be used for the development of an appropriate GRADE framework for public health policy guidelines.

## Methods

We conducted this systematic review following the preferred reporting items for systematic reviews and meta-analyses statement (PRISMA) [23] and the Cochrane Handbook for Systematic Reviews of Interventions [24].

### Inclusion and exclusion criteria

#### Types of studies

Randomized controlled trials (RCTs) run in healthcare settings were considered eligible. Randomization was allowed both at individual and cluster level.

#### Population

HWCs exposed to SARS-CoV-2 or any other respiratory infection. *Subgroups*: in-patient versus out-patient hospital setting.

#### Types of interventions

N95 respirators versus surgical masks.

#### Types of outcomes and assessment measures

We identified a priori the following outcomes, rated for importance as critical, important and not important.

- *Critical outcomes:* (i) SARS-CoV-2 infection; (ii) Clinical respiratory illness (CRI).
- *Important outcomes:* (iii) Influenza like illness (ILI); (iv) Laboratory-confirmed respiratory viral infection; (v) Laboratory-confirmed bacterial colonization; (vi) Laboratory-confirmed respiratory infection; (vii) Laboratory-confirmed influenza.
- *Not important outcome:* (viii) Discomfort of wearing respiratory protections. Outcome definitions are reported in S1 Appendix.

### Search strategy, study selection and data extraction

The full search strategy is reported in S1 Appendix.

### Risk of bias assessment

Two reviewers independently assessed the risk of bias (RoB) of the selected RCTs using the Cochrane Risk of Bias tool. Also, in cluster-RCTs specific risk of bias were considered. Further details are reported in S1 Appendix.

### Data Analysis and Synthesis of Results

We pooled data from studies with similar interventions and outcomes (for the intention-to-treat analysis) to calculate relative risk (RR) and the corresponding 95% confidence intervals (CIs). For cluster RCTs, we applied the specific method described in the Cochrane Handbook [25] to account for clustering and obtain adjusted RR and CIs. When examining critical outcomes we used the Claxton model for analyzing the value of the immediate implementation of an intervention versus the added benefit in reducing uncertainty derived from further research[26]. See S1 Appendix for details.

### Quality of the evidence-GRADE approach

We evaluated the overall quality of the evidence for critical and important outcomes using the GRADE approach[27]. Adjusted estimates were considered for judging the quality of the evidence. For critical outcomes, absolute effects were calculated at 95% CI and 90% CI[28]. A ‘summary of findings’ including the quality of the evidence, reasons for limitation and main findings were displayed in table. See S1 Appendix for details.

## Results

### Study selection

A total of 390 records resulted from the search on the electronic databases. Overall, we included four RCTs from five publications, of which one was an individual participants randomized trial [29] and three were cluster randomized trials [30-32]. One publication included additional outcomes related to one cluster RCTs [33]. Flow diagram of the study selection process is displayed in S1 Appendix.

### Description of the included studies

Overall, 8736 participants were considered, with the number of participants for each trial ranging from 446 to 5180. Three cluster randomized studies were performed in an inpatient [30-32] and one in an outpatient [34] setting.

**Table 1.**
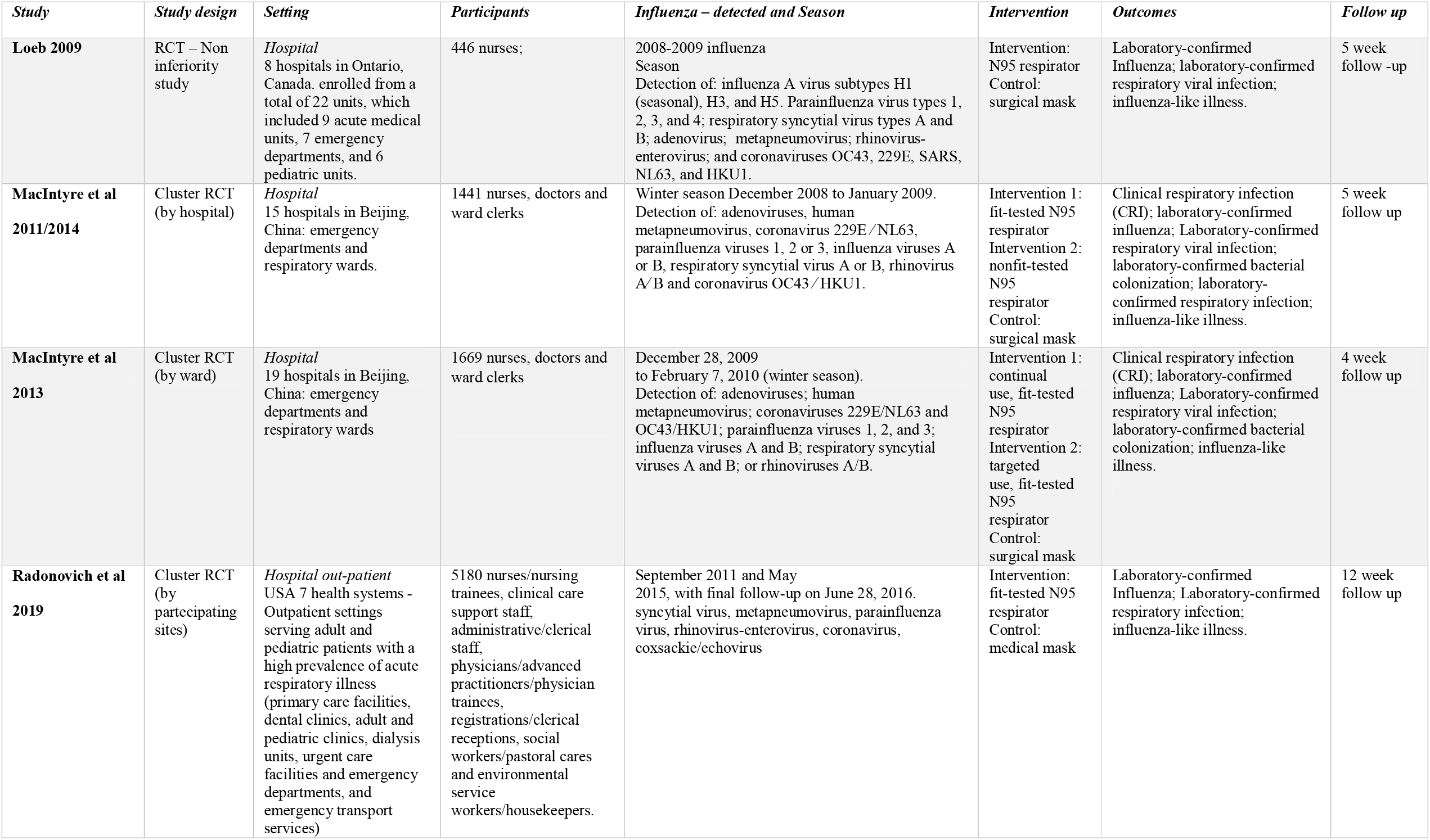
General Characteristics of included studies.

### Risk of bias

In the S1 Appendix we show the risk of bias of included studies: Loeb et al. 2009 [29] was judged at low risk of bias, the remaining [30-33] were assessed for additional bias related to clustering. Overall, four cluster randomized controlled trial were assessed as high risk of bias for imbalance at baseline.

### Critical Outcomes

No RCTs addressing the prevention of SARS-CoV-2 infection among HCWs was found. For CRI, we included two cluster RCTs with 2594 HCWs from in-patient hospital setting[30, 31]. Adjusting data for clustering, using N95 respirators reduced meaningfully the risk of developing CRI respect to surgical masks (2 RCTs, RR 0.43, 95% CI 0.29, 0.64; I^2^=0%) (Fig 1), with low quality of evidence and an absolute effect of preventing 73 more (95% CI from 91 more to 46 more) infections per 1000 HCWs wearing N95 respirators (Table 2). According to the Claxton model[26], in the worst case scenario the added benefit of more research in reducing uncertainty would be of reducing to 51 infections (upper 90% CI limit) prevented per 1000 HCWs wearing N95 respirators compared to surgical masks) (Fig 2).

**Table 2.**
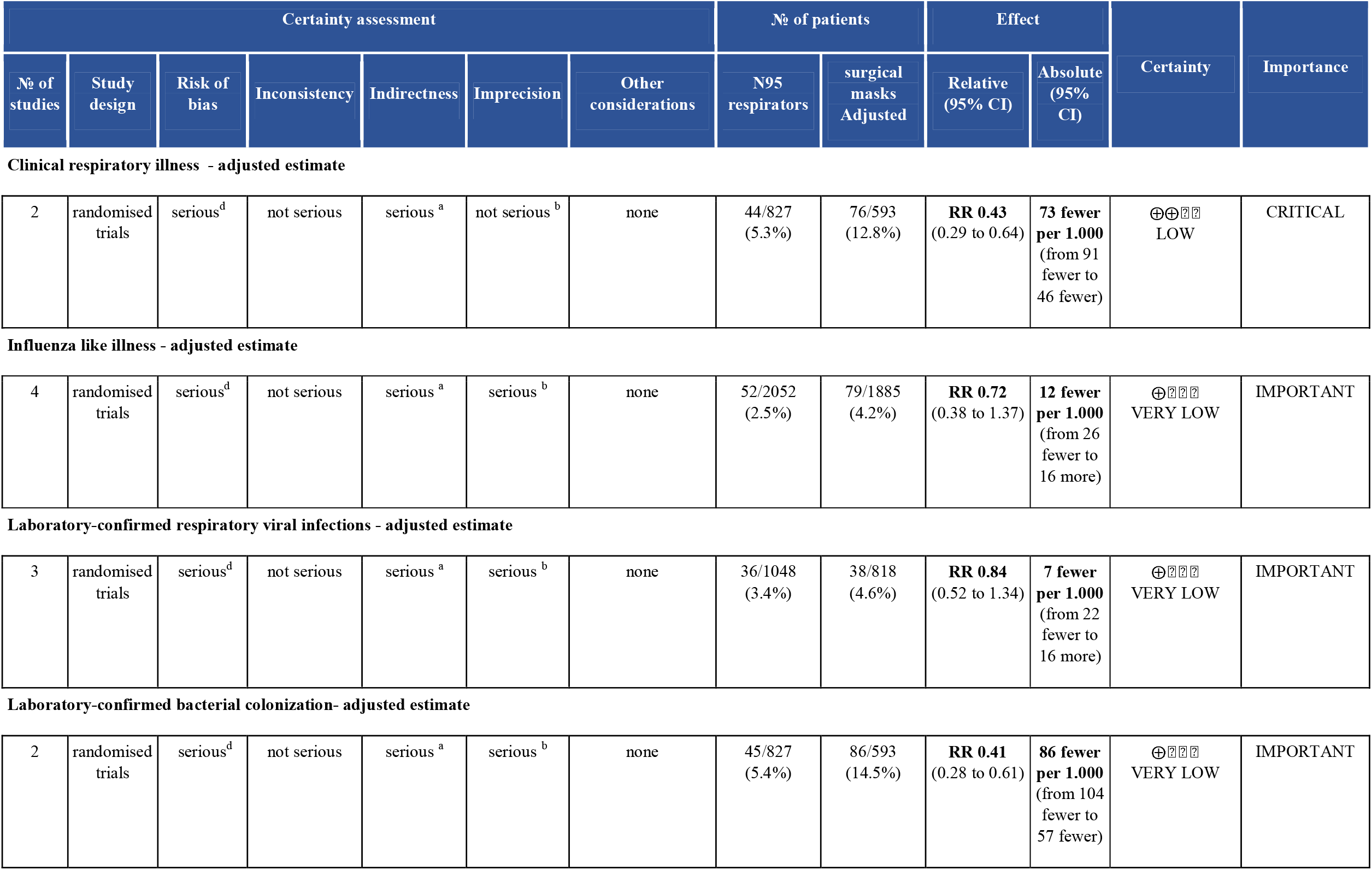
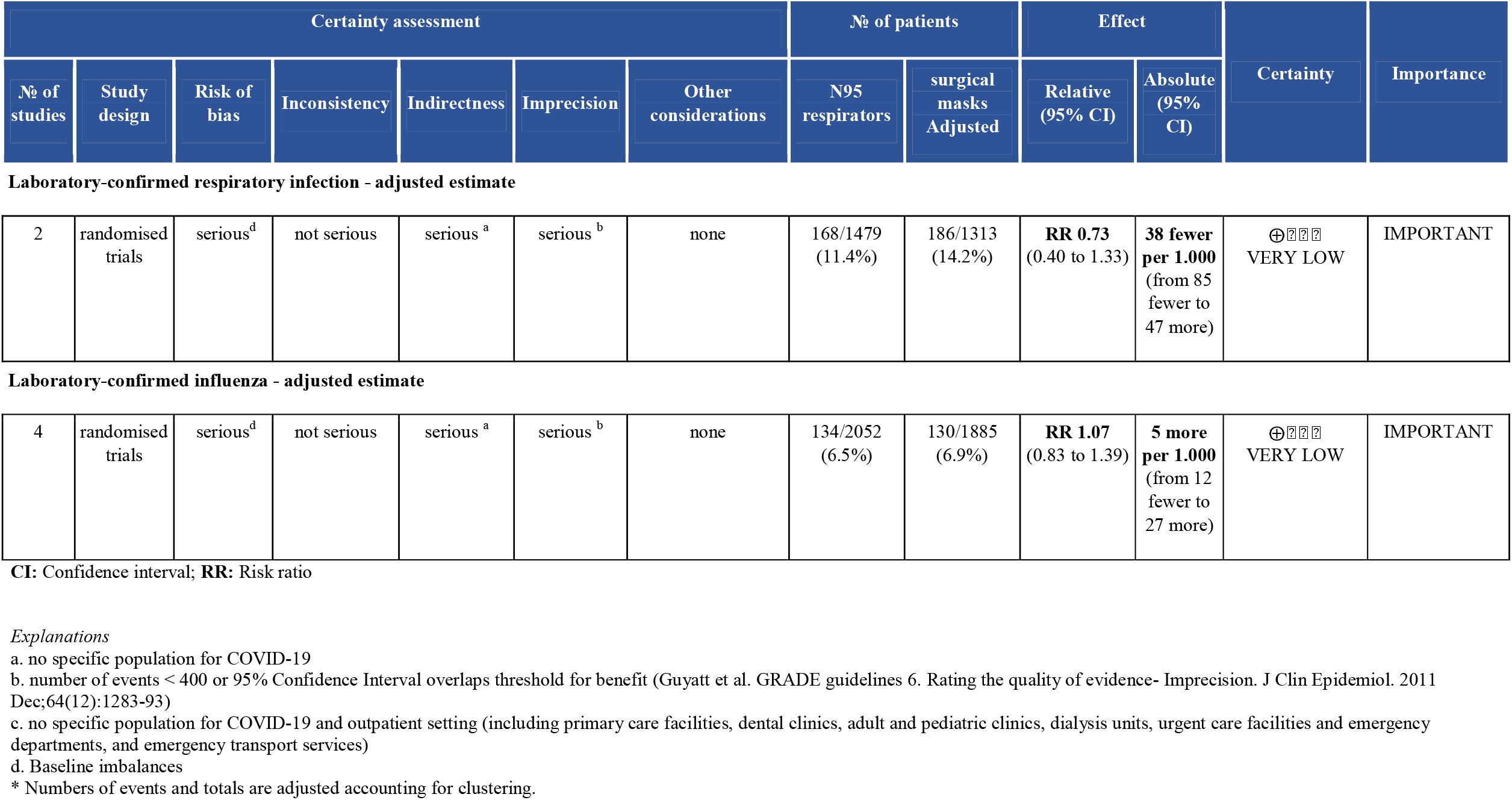
GRADE Summary of findings table.

**Fig 1.**
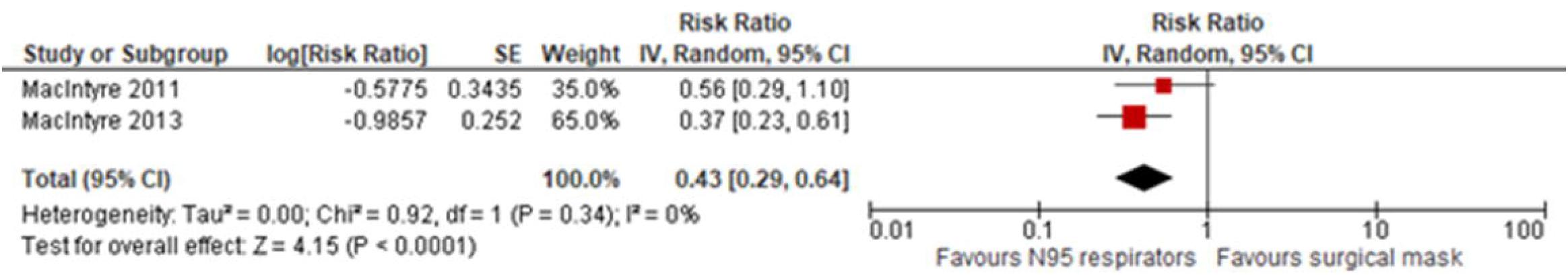
Forest plot of clinical respiratory illness (CRI) – random effect model meta-analysis with 95% CI.

**Fig 2.**
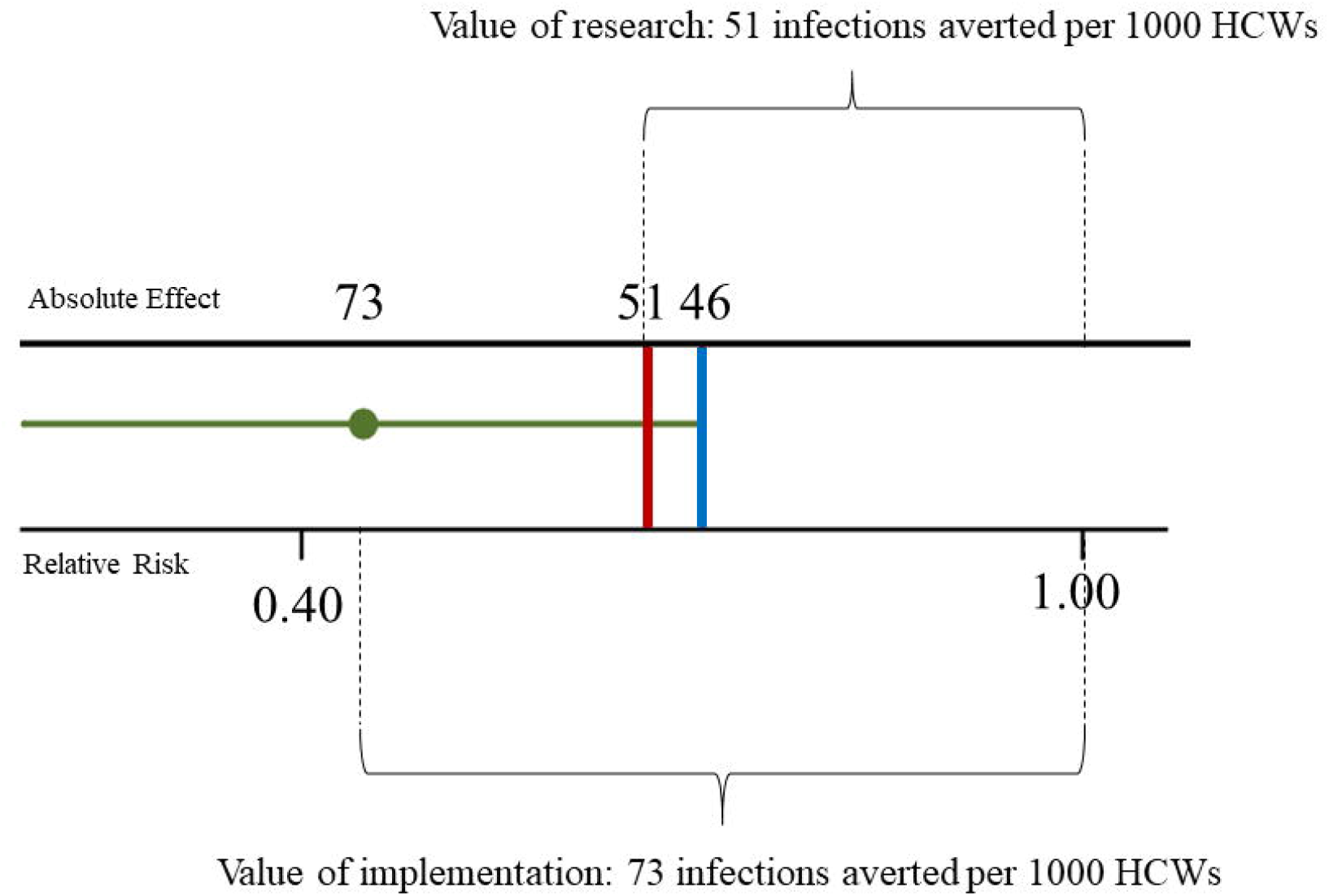
Trade-offs between implementation and deferral of the intervention about clinical respiratory. illness (CRI). The green line represents the 95% CI of the overall effect of N95 respirators respect to surgical masks for CRI. The blue line is the upper limit of 95% CI, RR 0.43 [0.29-0.64] anticipated absolute effect: 73 [91 46], the red line is the upper limit of 90% CI, RR 0.43 [0.31-0.60] anticipated absolute effect: 73 [88 51]. Value of implementation is therefore of 73 infections averted × 1000 HCW, value of further research is of 51 infections averted × 1000 HCW.

## Other outcomes

The quality for the evidence was very low for all the important outcomes (Table 2). A trend in favour of N95 was found for ILI (4 RCTs, 8220 HCWs; RR 0.72, 95% CI 0.38, 1.37; I^2^=24%), laboratory confirmed respiratory viral infections (3 RCTs, 3040 HCWs; RR 0.84, 95% CI 0.52, 1.34; I^2^=0%), laboratory confirmed respiratory infection (2 RCTs, 6221 HCWs; RR 0.73, 95% CI 0.40, 1.33; I^2^=69%), laboratory confirmed influenza (4 RCTs, 8220 HCWs; RR 1.07, 95% CI 0.83, 1.39; I^2^=0%). The protective effect of N95 respirators for bacterial respiratory colonisation was significant (2 RCTs, 2594 HCWs; RR 0.41, 95% CI 0.28, 0.61; I^2^=0%).

Discomfort (not important outcome) was higher among HCWs wearing N95 respirators [30]: data are reported through descriptive statistics in S1 Appendix.

## Discussion

The exceptional threat to the resilience of health care systems posed by this pandemic is well known and protecting HCWs is among the most important interventions for successfully managing the COVID-19 pandemic. There is no agreement among healthcare organisations on whether HCWs should wear surgical masks or N95 respirators during the routine care (not involving aerosol generating procedures) of COVID-19 suspected or affected patients [5-7].

While some observational evidence suggests that an airborne (aerosol) route of diffusion of SARS-CoV-2 may occur also outside the aerosol generating procedures (such as tracheal intubation, sputum induction and airway suctioning) so far no RCT has directly compared the effect of filtering respirators with surgical masks in preventing SARS-CoV-2 infection among HCWs. Also, it is unlikely that such a trial could be ethically acceptable in the near future if the evidence of an aerosol diffusion of SARS-CoV-2 grows even more. Indeed, in our meta-analysis we found that wearing N95 halves the risk of any clinical respiratory infection in HCWs compared to wearing only surgical masks. An immediate implementation of the intervention (wearing N95 respirator) could avoid 73 respiratory infections per 1000 HCWs. According to the Claxton model[26], the added value of further research in reducing uncertainty would be to avoid no more than 51 infections per 1000 HCW, in the worst case scenario, making deferral of this intervention while awaiting more studies unreasonable.

The favourable (albeit not significant) trend of N95 for laboratory confirmed respiratory infections and ILI deserves some comment. In fact, these findings could be viewed as evidence against the benefit of respirators. Instead, given the blurred distinction between airborne and droplet diffusion of respiratory virues[35] it could also be considered as indirect evidence of the opportunistic airborne route of transmission of respiratory viruses in the healthcare environment, where prolonged exposures, high viral loads, asymptomatic carriers, overcrowding and poor ventilation could enhance the opportunistic airborne diffusion among HCWs of viruses such as SARS-CoV-2. Regarding the lack of apparent benefit of N95 for influenza in the only trial where this outcome was assessed, both the outpatient setting (with lower viral exposure loads)[36] and the droplet route of transmission believed to be operative for influenza are worth of consideration. Finally, we suggest to integrate the perspective and the findings of this review into the appropriate GRADE framework, considering the added difficulties of urgency and uncertainty, which make the production of a reliable guideline even more challenging [37]. Such guidelines should explicitly consider among other factors the human and organizational costs of delaying the adoption of N95 respirators versus the benefits of an immediate adoption and, finally, the key value of safeguarding HCWs in the context of SARS-CoV-2 pandemic.

## Limitations and strengths

Several limitations should be considered. Wearing N95 respirators is only one component among a series of complex procedures, so that the identified effect cannot exclusively be attributed to this intervention. The source of infection (community rather than the workplace) cannot be ascertained in any of the trials. Second, one RCT required HCWs to wear N95 respirators only when caring for patients with febrile respiratory illness[29], whereas all others specified continuous respirator use. Third, one study was performed in an outpatient setting that can be considered at moderate risk of transmission[32]. Finally, our meta-analyses did not investigate the adherence of wearing an N95 respirator. One of the included trials reported discomfort of using N95 respirators[30].

The main strength of our study was the use of appropriate Cochrane methods for analysing cluster randomized studies. By inflating variances this method allows to obtain adjusted estimates of relative risks. Indeed, if clustering is ignored, P values will be artificially small resulting in false positive conclusions about the effectiveness of the intervention. In addition, we adopted the Claxton model to quantify the trade-off between immediate implementation of the intervention versus deferring it while awaiting further evidence. Finally, we offered clinical recommendations based on the quality of evidence by GRADE approach[38].

## Conclusion

This is the first systematic review on the efficacy of N95 respirators versus surgical masks among HCWs accounting for possible bias derived from cluster trials and evaluating the findings from a public health policy perspective. We found evidence that N95 respirators halve the risk of any respiratory infection compared to surgical masks. Considering that the absenteeism from work due to healthcare related infections hampers heavily the resilience of healthcare systems facing an infectious pandemic, the protective effect of N95 respirators for this critical outcome could produce large benefits in the current context. Furthermore, the immediate implementation of the intervention, rather than deferring it until more studies will be available, seems justified on a sound quantitative basis. The evidence from the current study could be used to inform the production of trustworthy GRADE based guidelines for the prevention of SARS-CoV-2 infection among HCWs.

## Data Availability

All data are included in the manuscript.

## Declaration of interests

All authors declare no competing interests.

## Funding

None.

GC confirm that she had full access to all the data in the study and had final responsibility for the decision to submit for publication.

## Notes

### Competing Interest Statement

The authors have declared no competing interest.

## Bibliography

1. WHO Director-General’s opening remarks at the media briefing on COVID-19—11 March 2020. World Health Organization. 11 March 2020. Accessed on march 29 2020 [Available from: https://www.who.int/dg/speeches/detail/who-director-general-s-opening-remarks-at-the-media-briefing-on-covid-1911-march-2020].

2. Coronavirus disease 2019 (COVID-19): situation report-70. Published March 30, 2020. Accessed March 18, 2020. https://www.who.int/emergencies/diseases/novel-coronavirus-2019/situation-reports.

3. Riccardo F, Andrianou X, Bella A, et al. Integrated surveillance of COVID-19 in Italy. Accessed on March 29 2020 [Available from: https://www.epicentro.iss.it/coronavirus/bollettino/Infografica_30marzo%20ENG.pdf.

4. https://portale.fnomceo.it/elenco-dei-medici-caduti-nel-corso-dellepidemia-di-covid-19/.

5. World Health Organisation (WHO), 2020. Novel Coronavirus (2019-nCoV).

6. European Centre for Disease Prevention and Control (ECDC), 2020. Infection Prevention and Control For the Care of Patients With 2019-nCoV in Healthcare Settings. Accessed on march 27 2020. [Available from: https://www.ecdc.europa.eu/sites/default/files/documents/nove-coronavirus-infection-prevention-control-patients-healthcare-settings.pdf.

7. Center for Disease Control and Prevention (CDC), 2020. Interim Healthcare Infection Prevention and Control Recommendations For Patients Under Investigation for 2019 Novel Coronavirus January 2020. Accessed on march 27 2020.[Available from: https://www.cdc.gov/coronavirus/2019-nCoV/infection-control.html.

8. Cheng VCC, Wong SC, Chen JHK, Yip CCY, Chuang VWM, Tsang OTY, et al. Escalating infection control response to the rapidly evolving epidemiology of the Coronavirus disease 2019 (COVID-19) due to SARS-CoV-2 in Hong Kong. Infect Control Hosp Epidemiol. 2020:1–24.

9. Ong SWX, Tan YK, Chia PY, Lee TH, Ng OT, Wong MSY, et al. Air, Surface Environmental, and Personal Protective Equipment Contamination by Severe Acute Respiratory Syndrome Coronavirus 2 (SARS-CoV-2) From a Symptomatic Patient. JAMA. 2020.

10. World Health Organization. Report of the WHO-China Joint Mission on Coronavirus Disease 2019 (COVID-19) 16-24 February 2020 [Internet]. Geneva: World Health Organization; 2020.

11. Schwartz KL, Murti M, Finkelstein M, Leis J, Fitzgerald-Husek A, Bourns L, et al. Lack of COVID-19 Transmission on an International Flight. CMAJ. Published on: 24 February 2020 https://www.cmaj.ca/content/192/7/E171/tab-e-letters#lack-of-covid-19-transmission-on-aninternational-flight.

12. McKinney. K.R., Gong. Y.Y.T.G. L. Environmental transmission of SARS at amoy gardens. J Environ Health 68 (9), 26–30 quiz 51-2 2006.

13. Chughtai AA, Seale H, Islam MS, Owais M, Macintyre CR. Policies on the use of respiratory protection for hospital health workers to protect from coronavirus disease (COVID-19). International journal of nursing studies. 2020;105:103567.

14. van Doremalen N, Bushmaker T, Morris DH, Holbrook MG, Gamble A, Williamson BN, et al. Aerosol and Surface Stability of SARS-CoV-2 as Compared with SARS-CoV-1. The New England journal of medicine. 2020.

15. Joshua L Santarpia DNR, Vicki Herrera, M. Jane Morwitzer, Hannah Creager, George W. Santarpia, Kevin K Crown, David Brett-Major, Elizabeth Schnaubelt, M. Jana Broadhurst, James V. Lawler, St. Patrick Reid, John J. Lowe. Transmission Potential of SARS-CoV-2 in Viral Shedding Observed at the University of Nebraska Medical Center. 2020.

16. Lindsley WG, King WP, Thewlis RE, Reynolds JS, Panday K, Cao G, et al. Dispersion and exposure to a cough-generated aerosol in a simulated medical examination room. Journal of occupational and environmental hygiene. 2012;9(12):681–90.

17. Lindsley WG, Blachere FM, Thewlis RE, Vishnu A, Davis KA, Cao G, et al. Measurements of airborne influenza virus in aerosol particles from human coughs. PloS one. 2010;5(11):e15100.

18. Long Y, Hu T, Liu L, Chen R, Guo Q, Yang L, et al. Effectiveness of N95 respirators versus surgical masks against influenza: A systematic review and meta-analysis. Journal of evidence-based medicine. 2020.

19. Offeddu V, Yung CF, Low MSF, Tam CC. Effectiveness of Masks and Respirators Against Respiratory Infections in Healthcare Workers: A Systematic Review and Meta-Analysis. Clinical infectious diseases : an official publication of the Infectious Diseases Society of America. 2017;65(11):1934–42.

20. Cowling BJ, Zhou Y, Ip DK, Leung GM, Aiello AE. Face masks to prevent transmission of influenza virus: a systematic review. Epidemiology and infection. 2010;138(4):449–56.

21. Remuzzi A, Remuzzi G. COVID-19 and Italy: what next? Lancet. 2020.

22. Guyatt G. GRADE weak or conditional recommendations mandate shared decision-making. Author Response. J Clin Epidemiol. 2018;102:147–8.

23. Moher D, Liberati A, Tetzlaff J, Altman DG, Group P. Preferred reporting items for systematic reviews and meta-analyses: the PRISMA statement. BMJ. 2009;339:b2535.

24. Higgins JPT GS. Cochrane Handbook for Systematic Reviews of Interventions. Version 5.1.0 [updated March 2011]. The Cochrane Collaboration, 2011. Available from www.handbook.cochrane.org. 2011. Available from: www.handbook.cochrane.org

25. Higgins JPT GS. Cochrane Handbook for Systematic Reviews of Interventions. Chapter 16: Special topics in statistics. Version 5.1.0 [updated March 2011]. The Cochrane Collaboration, 2011. [Available from: https://handbook-5-1.cochrane.org/chapter_16/16_3_2_assessing_risk_of_bias_in_cluster_randomized_trials.htm.

26. Claxton K, Griffin S, Koffijberg H, McKenna C. How to estimate the health benefits of additional research and changing clinical practice. BMJ. 2015;351:h5987.

27. Guyatt G, Oxman AD, Akl EA, Kunz R, Vist G, Brozek J, et al. GRADE guidelines: 1. Introduction-GRADE evidence profiles and summary of findings tables. J Clin Epidemiol. 2011;64(4):383–94.

28. Guyatt GH, Oxman AD, Santesso N, Helfand M, Vist G, Kunz R, et al. GRADE guidelines: 12. Preparing summary of findings tables-binary outcomes. J Clin Epidemiol. 2013;66(2):158–72.

29. Loeb M, Dafoe N, Mahony J, John M, Sarabia A, Glavin V, et al. Surgical mask vs N95 respirator for preventing influenza among health care workers: a randomized trial. Jama. 2009;302(17):1865–71.

30. MacIntyre CR, Wang Q, Cauchemez S, Seale H, Dwyer DE, Yang P, et al. A cluster randomized clinical trial comparing fit-tested and non-fit-tested N95 respirators to medical masks to prevent respiratory virus infection in health care workers. Influenza Other Respir Viruses. 2011;5(3):170–9.

31. MacIntyre CR, Wang Q, Seale H, Yang P, Shi W, Gao Z, et al. Arandomized clinical trial of three options for N95 respirators and medical masks in health workers. Am J Respir Crit Care Med. 2013;187(9):960–6.

32. Radonovich LJ, Jr., Simberkoff MS, Bessesen MT, Brown AC, Cummings DAT, Gaydos CA, et al. N95 Respirators vs Medical Masks for Preventing Influenza Among Health Care Personnel: A Randomized Clinical Trial. Jama. 2019;322(9):824–33.

33. MacIntyre CR, Wang Q, Rahman B, Seale H, Ridda I, Gao Z, et al. Efficacy of face masks and respirators in preventing upper respiratory tract bacterial colonization and co-infection in hospital healthcare workers. Prev Med. 2014;62:1–7.

34. MacIntyre CR, Cauchemez S, Dwyer DE, Seale H, Cheung P, Browne G, et al. Face mask use and control of respiratory virus transmission in households. Emerg Infect Dis. 2009;15(2):233–41.

35. Tellier R, Li Y, Cowling BJ, Tang JW. Recognition of aerosol transmission of infectious agents: a commentary. BMC Infect Dis. 2019;19(1):101.

36. McDiarmid M, Harrison R, Nicas M. N95 Respirators vs Medical Masks in Outpatient Settings. JAMA. 2020;323(8):789.

37. Moberg J, Oxman AD, Rosenbaum S, Schunemann HJ, Guyatt G, Flottorp S, et al. The GRADE Evidence to Decision (EtD) framework for health system and public health decisions. Health Res Policy Syst. 2018;16(1):45.

38. Andrews J, Guyatt G, Oxman AD, Alderson P, Dahm P, Falck-Ytter Y, et al. GRADE guidelines: 14. Going from evidence to recommendations: the significance and presentation of recommendations. J Clin Epidemiol. 2013;66(7):719–25.

